# Pharmacovigilance Analysis on Cerebrovascular Accidents and Coronavirus disease 2019 Vaccines

**DOI:** 10.1101/2021.04.19.21255768

**Authors:** Lt. Pushkar Aggarwal

## Abstract

**Introduction:** Recently, there have been reports of cerebrovascular accidents (CVA) occurring in individuals who have received the Coronavirus disease 2019 (COVID-19) vaccine.

**Objective:** The objective of this analysis was to determine if a statistically significant signal exists in post-marketing safety reports between CVA and the three COVID-19 vaccines being administered in the United States of America (Pfizer, Moderna, Janssen).

**Methods:** A pharmacovigilance disproportionality analysis on adverse events reported with COVID-19 vaccines was conducted using data from Vaccine Adverse Event Reporting System.

**Results:** A statistically significant signal was found between CVA events and each of the three COVID-19 vaccines (Pfizer/BioNTech’s, Moderna’s and Janssen’s) in the VAERS database. Females and individuals of age 65 or older had higher number of case reports of CVA events with the COVID-19 vaccines. Females had also more COVID-19 adverse event reports in which a CVA was reported and resulted in the patient having permanent disability or death.

**Limitations:** Randomized controlled trials are needed to further analyze this signal.

**Conclusion:** Patients should be made aware of the risk-benefit and symptoms to watch out for that may indicate the onset of a CVA and informed to seek medical care as soon as possible if they develop these symptoms.

## Introduction

Cerebrovascular accident (CVA), also known as acute strokes, results in the loss of blood flow, nutrients and oxygen to a region of the brain.^1^ This causes neuronal damage and can lead to permanent neurological deficits. Cerebrovascular accidents are broadly classified into ischemic or hemorrhagic etiologies. Some causes of CVA include uncontrolled hypertension, development of arteriosclerosis, emboli and hypercoagulable state.

Recently, there have been reports of CVA occurring in individuals who have received the Coronavirus disease 2019 (COVID-19) vaccine. The objective of this analysis was to determine if a statistically significant signal exists in post-marketing safety reports between CVA and the three COVID-19 vaccines being administered in the United States of America (Pfizer, Moderna, Janssen).

## Methods

Adverse events/reaction case reports from vaccines were acquired from the Vaccine Adverse Event Reporting System (VAERS).^2^ VAERS is a database co-managed by the Centers for Disease Control and Prevention (CDC) and the U.S. Food and Drug Administration. The database contains adverse events/reaction case reports from vaccines that are submitted voluntarily by healthcare providers, vaccine manufacturers, and the public from 1990 to present. For COVID-19 vaccination adverse events, healthcare providers are required to report adverse events that resulted in death, life-threatening adverse event and permanent disability.

All adverse events/reaction case reports from January 1, 1990 to April 10, 2021 were acquired from VAERS. These were stratified by the adverse event “Cerebrovascular Accident” reported to be associated with any vaccine. They were further examined as being associated with COVID-19 vaccines and by COVID-19 vaccine manufacturers (Janssen, Moderna or Pfizer/BioNTech). In addition, demographic factors (gender, age) and the severity of the adverse event (death, life-threatening, permanent disability) were analyzed.

A pharmacovigilance disproportionality analysis on adverse events reported with COVID-19 vaccines was conducted using data from VAERS. A disproportionality analysis compares the expected count to the observed count for a vaccine and adverse event combination. This allows for the determination of whether a statistically significant signal exists between the vaccine and adverse event. The disproportionality analysis includes relative reporting ratio (RRR), proportional reporting ratio (PRR), reporting odds ratio (ROR) and chi squared with Yates’ correction.

Other disproportionality measures which can be used to decipher a signal are information component (IC) and the empirical Bayes geometric mean (EBGM). These algorithms, however, differ from the above disproportionality algorithms in that the PRR, RRR and ROR utilize a frequentist approach, whereas the IC and EBGM utilize a Bayesian approach.^3^ The PRR is currently used by the UK Medicines and Healthcare products Regulatory Agency (MHRA), the ROR by the Netherlands Pharmacovigilance Centre and the IC by the World Health Organization (WHO). The Bayesian Confidence Propagation Neural Network (BCPNN) analysis was proposed based on Bayesian logic where the relation between the prior and posterior probability was expressed as the information component. The IC given by the BCPNN is applied by the WHO Uppsala Monitoring Center (UMC). Signal detection using IC is done using the IC_025_ metric, a criterion indicating the lower bound of the 95% two-sided confidence interval of the IC, and a signal is detected if the IC_025_ value exceeds zero.^4^ Evans et al. define a criterion by which to evaluate whether the disproportionality analysis indicates a statistically significant signal or not: a PRR greater than or equal to 2, chi-squared greater than or equal to 4 and number of events greater than or equal to 3.^5^ In addition, according to Van Puijenbroek et al., a lower 95% CI of ROR greater than 1 indicates a statistically significant signal between a drug and an event.^6^ In this analysis, the adverse event signal is reported using frequentist methods of RRR, ROR and PRR and the Bayesian based IC_025_ metric.^7^

Approval by institutional review board, or human subjects’ committee was not required as the analysis was performed on de-identified retrospective public domain safety data.

## Results

A total of 31,459 adverse events with Pfizer/BioNTech’s vaccine were reported in the VAERS database. Of these, 165 had a CVA event (0.52% of reported events) and 27 of the 165 reports resulted in death. A total of 29,913 adverse events with Moderna’s vaccine were reported. Of these, 145 had a CVA event (0.48% of reported events) and 27 of the 145 reports resulted in death. A total of 6,751 adverse events with Janssen’s vaccine were reported. Of these, 17 had a CVA event (0.25% of reported events) with 0 of the 17 reports resulting in death (Table 1).

**Table 1.**
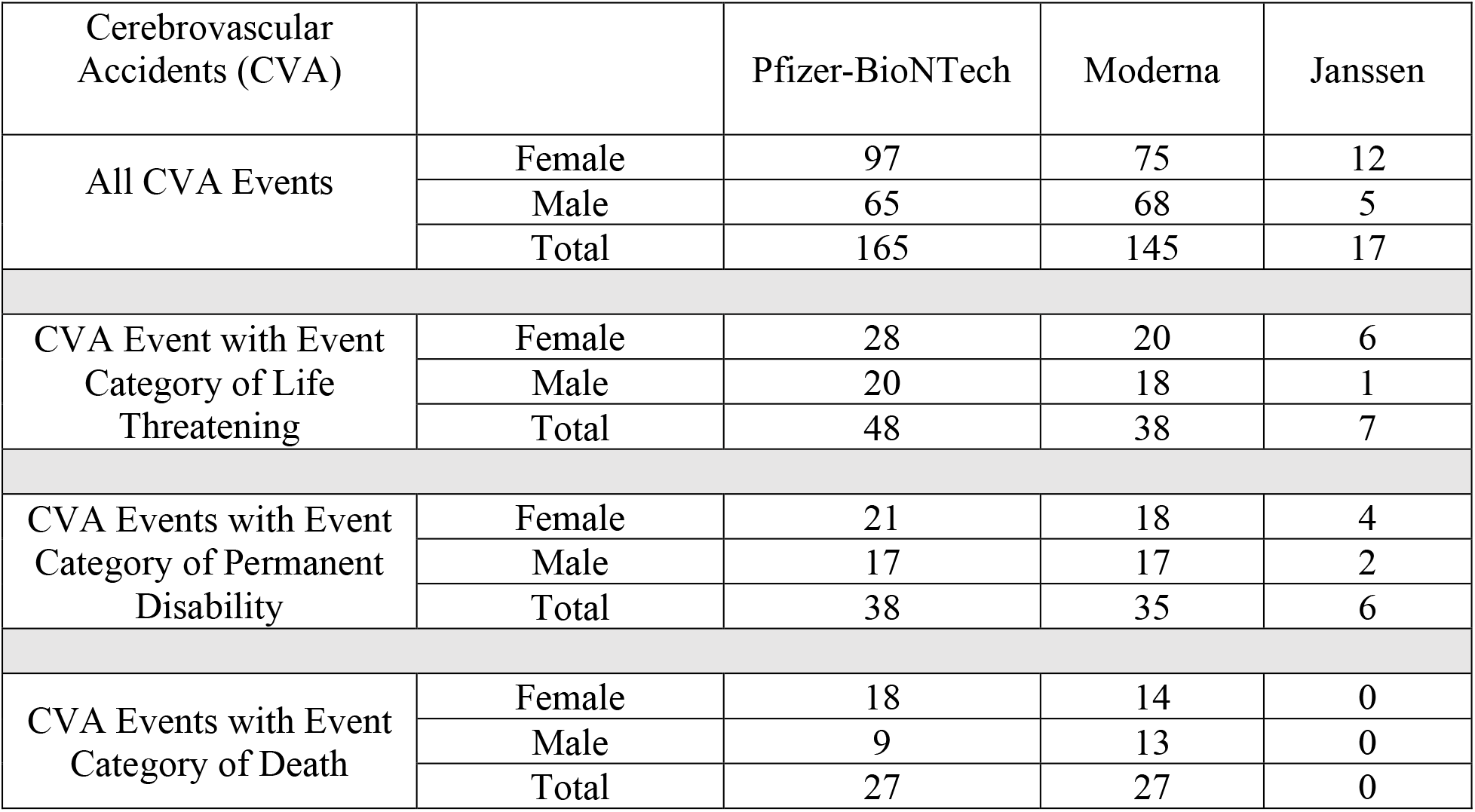
Cerebrovascular Accidents (CVA) by COVID-19 Vaccine Stratified by Gender and by Event Category

### Stratification by Gender

Females had a higher number of reports of CVA events reported with each of the three COVID-19 vaccines than males. In addition, females had more reports of life threatening CVA events and permanent disability with a CVA with each of the three COVID-19 vaccines than males. Females had more reports of death and a CVA with both Pfizer/BioNTech’s vaccine and Moderna’s vaccine (Table 1).

### Stratification by Age

The majority of CVA events with the COVID-19 vaccines occurred in the age group 65 or older. In the age group of 65 or older, Pfizer/BioNTech’s vaccine had 111 CVA reports, Moderna’s vaccine had 114 CVA reports and Janssen’s vaccine had 9 CVA reports (Table 2).

**Table 2.** Cerebrovascular Accidents (CVA) by COVID-19 Vaccine Stratified by Age

|  | Age (years of age) | Pfizer-BioNTech | Moderna | Janssen |
| --- | --- | --- | --- | --- |
| CVA Events | 18-29 | 2 | 1 | 0 |
|  | 30-39 | 8 | 1 | 0 |
|  | 40-49 | 10 | 4 | 1 |
|  | 50-59 | 12 | 10 | 3 |
|  | 60-64 | 6 | 7 | 1 |
|  | 65+ | 111 | 114 | 9 |
|  | Unknown | 16 | 8 | 3 |
|  | Total | 165 | 145 | 17 |

### Statistical Analysis

For CVA events with Pfizer/BioNTech’s vaccine, the PRR was 6.00, lower 95% ROR was 5.08, chi-square with Yates’ correction was 544.31, and IC_025_ was 1.91 (statistically significant signal) (Table 3). For CVA events with Moderna’s vaccine, the PRR was 5.39, lower 95% ROR was 4.52, chi-square with Yates’ correction was 422.66, and IC_025_ was 2.04 (statistically significant signal). For CVA events with Janssen’s vaccine, the PRR was 2.42, lower 95% ROR was 1.50, chi-square with Yates’ correction was 12.55, and IC_025_ was 0.39 (statistically significant signal). Disproportionality analysis results by gender and for age greater than or equal to 65 are provided in Table 3.

**Table 3.**
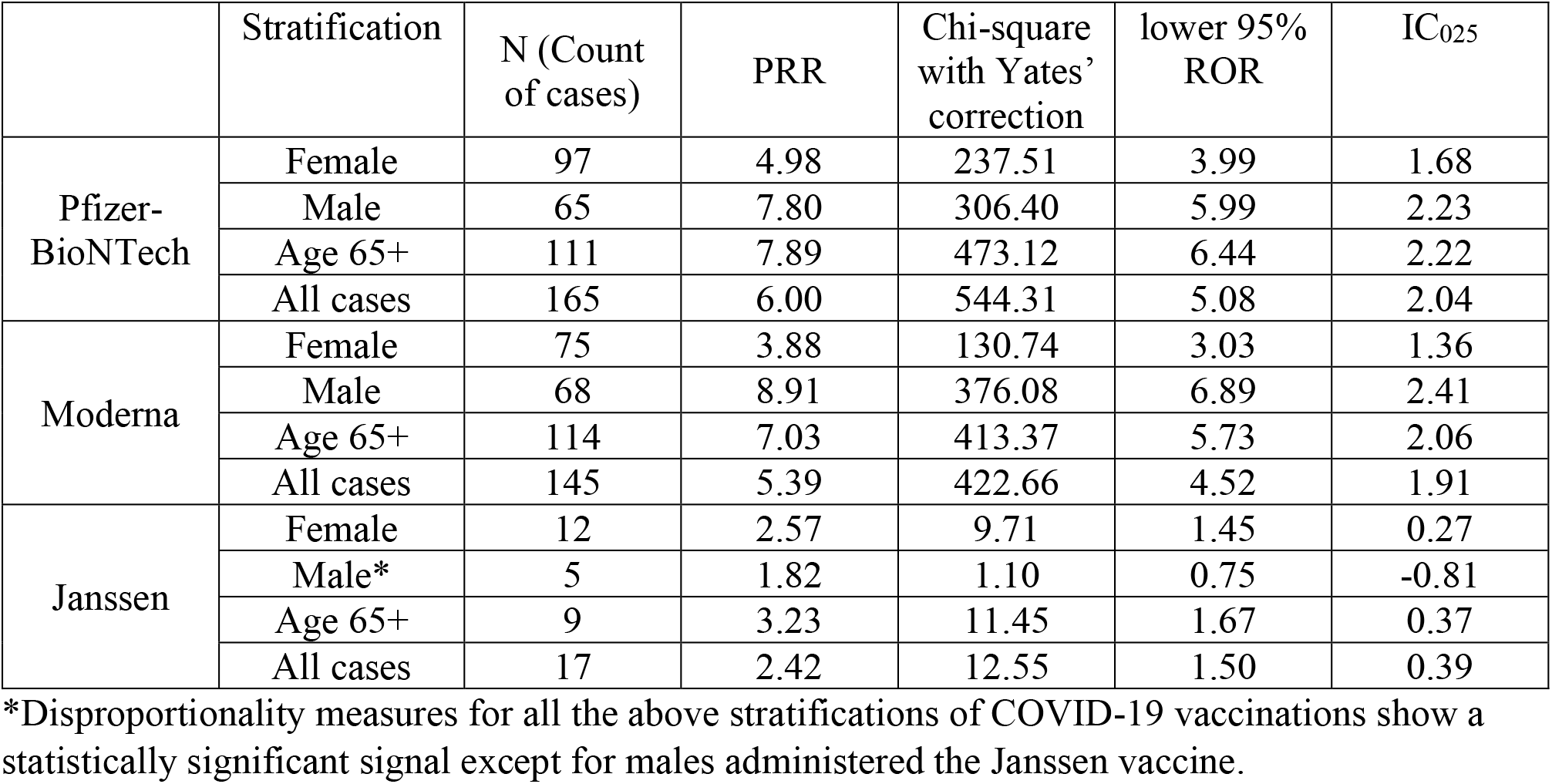
Disproportionality Analysis for COVID-19 Vaccinations and Cerebrovascular Accidents

|  | Stratification | N (Count of cases) | PRR | Chi-square with Yates' correction | lower 95% ROR | IC<sub>025</sub> |
| --- | --- | --- | --- | --- | --- | --- |
| Pfizer-BioNTech | Female | 97 | 4.98 | 237.51 | 3.99 | 1.68 |
|  | Male | 65 | 7.80 | 306.40 | 5.99 | 2.23 |
|  | Age 65+ | 111 | 7.89 | 473.12 | 6.44 | 2.22 |
|  | All cases | 165 | 6.00 | 544.31 | 5.08 | 2.04 |
| Moderna | Female | 75 | 3.88 | 130.74 | 3.03 | 1.36 |
|  | Male | 68 | 8.91 | 376.08 | 6.89 | 2.41 |
|  | Age 65+ | 114 | 7.03 | 413.37 | 5.73 | 2.06 |
|  | All cases | 145 | 5.39 | 422.66 | 4.52 | 1.91 |
| Janssen | Female | 12 | 2.57 | 9.71 | 1.45 | 0.27 |
|  | Male* | 5 | 1.82 | 1.10 | 0.75 | -0.81 |
|  | Age 65+ | 9 | 3.23 | 11.45 | 1.67 | 0.37 |
|  | All cases | 17 | 2.42 | 12.55 | 1.50 | 0.39 |

\*Disproportionality measures for all the above stratifications of COVID-19 vaccinations show a statistically significant signal except for males administered the Janssen vaccine.

## Discussion

A statistically significant signal was found between CVA events and each of the three COVID-19 vaccines (Pfizer/BioNTech’s, Moderna’s and Janssen’s) in the VAERS database. Females and individuals of age 65 or older had more case reports of CVA events with the COVID-19 vaccines. Females had also more COVID-19 adverse event reports in which a CVA was reported and resulted in the patient having permanent disability or death. Randomized controlled trials are needed to further analyze this signal. Patients should be made aware of the risk-benefit and symptoms to watch out for that may indicate the onset of a CVA and informed to seek medical care as soon as possible if they develop these symptoms.

As per VAERS, the adverse data reports in VAERS cannot be used to determine the rate/incidence of the adverse events. As of April 12, 2021, 97.9 million doses of Pfizer/BioNTech’s COVID-19 vaccine, 84.7 million doses of Moderna’s COVID-19 vaccine and 6.86 million doses of Janssen’s COVID-19 vaccine had been administered.^8^ As of April 10, 2021, 117 million people in US had been vaccinated for Covid-19.^9^ In 2011, ∼611,000 people in the United States experienced a first stroke, putting the incidence of stroke at 0.20%. The strokes caused ∼1 of every 20 deaths in the United States.^10^ Patients should be made aware of symptoms to watch out for that may indicate the onset of a CVA and informed to seek medical care as soon as possible if they develop these symptoms.

VAERS is a passive reporting system, meaning it relies on individuals to send in reports of their experiences to CDC and FDA. VAERS is not designed to determine if a vaccine caused a health problem, but is especially useful for detecting unusual or unexpected patterns of adverse event reporting that might indicate a possible safety problem with a vaccine. This way, VAERS can provide CDC and FDA with valuable information that additional work and evaluation is necessary to further assess a possible safety concern.

Not every adverse event is reported to VAERS, and therefore, the incidence of an adverse event cannot be calculated. However, if it is assumed that the database does not contain the unreported cases, it is likely that the incidence rates derived from the database will be lower than actual. These data and the analysis do not, by themselves, demonstrate causal associations. Randomized controlled studies are needed in order to establish causality. However, VAERS has advantages in that it can and has been used to identify vaccine safety signals in real-world situations, which is near impossible with the limited number of subjects used in the randomized clinical trials.^7^

## Data Availability

Data used for this analysis is publicly available.

## Acknowledgements

None

## References

1. Tadi P, Lui F. Acute Stroke. In: StatPearls. StatPearls Publishing; 2021. Accessed April 18, 2021. http://www.ncbi.nlm.nih.gov/books/NBK535369/

2. CDC and FDA. Vaccine Adverse Event Reporting System (VAERS). https://vaers.hhs.gov/

3. Tamura T, Sakaeda T, Kadoyama K, Okuno Y. Omeprazole- and Esomeprazole-associated Hypomagnesaemia: Data Mining of the Public Version of the FDA Adverse Event Reporting System. Int J Med Sci. 2012;9(5):322–326. doi:10.7150/ijms.4397

4. Bate A, Lindquist M, Edwards IR, et al. A Bayesian neural network method for adverse drug reaction signal generation. Eur J Clin Pharmacol. 1998;54(4):315–321. doi:10.1007/s002280050466

5. Evans SJ, Waller PC, Davis S. Use of proportional reporting ratios (PRRs) for signal generation from spontaneous adverse drug reaction reports. Pharmacoepidemiol Drug Saf. 2001;10(6):483–486. doi:10.1002/pds.677

6. van Puijenbroek EP, van Grootheest K, Diemont WL, Leufkens HG, Egberts AC. Determinants of signal selection in a spontaneous reporting system for adverse drug reactions. Br J Clin Pharmacol. 2001;52(5):579–586. doi:10.1046/j.0306-5251.2001.01501.x

7. Aggarwal P. Risk of bronchospasm and coronary arteriospasm with sugammadex use: a post marketing analysis. Ther Adv Drug Saf. 2019;10. doi:10.1177/2042098619869077

8. Tom Shimabukuro. Reports of cerebral venous sinus thrombosis with thrombocytopenia after Janssen COVID-19 vaccine. Presented at the: CDC COVID-19 Vaccine Task Force Vaccine Safety Team; April 14, 2021. https://www.cdc.gov/vaccines/acip/meetings/downloads/slides-2021-04/03-COVID-Shimabukuro-508.pdf

9. Roser M, Ritchie H, Ortiz-Ospina E, Hasell J. Coronavirus Pandemic (COVID-19). Our World Data. Published online March 5, 2020. Accessed April 18, 2021. https://ourworldindata.org/coronavirus

10. Mozaffarian D, Benjamin EJ, Go AS, et al. Heart disease and stroke statistics--2015 update: a report from the American Heart Association. Circulation. 2015;131(4):e29–322. doi:10.1161/CIR.0000000000000152

